# Career Intentions and Research-Environment Factors Across the Physician-Scientist Pipeline

**DOI:** 10.64898/2026.09.12.26362877

**Authors:** Sanaea Z. Bhagwagar, Abdelrahman Abushouk, John Dempsey, Daniel C. Brock, Aleksandar Obradovic, Alisha Faraz, James L Cross, Aisha Siebert, Han Naung Tun, Kevin M. Lin, Elena M. Wilson, Cynthia Y Tang, Auyon Ghosh, Mytien Nguyen, Alokkumar Jha, Daniel Shalev, Evan Noch, Jennifer M. Kwan

**Affiliations:** MD-PhD Program, State University of New York Upstate Medical University, Syracuse, NY, USA; Division of Cardiovascular Medicine, Brigham and Women’s Hospital, Harvard Medical School, Boston, MA, USA; Norton College of Medicine, State University of New York Upstate Medical University, Syracuse, NY, USA; Medical Scientist Training Program, Baylor College of Medicine, Houston, TX, USA; Department of Medicine, Columbia University, New York, NY, USA; Khan Lab school, Mountain View, CA, USA; Yale School of Medicine, New Haven, CT, USA; Department of Pediatric Urology and Medical Genetics & Genomics, Boston Children’s Hospital, Boston, MA, USA; Geisel School of Medicine at Dartmouth College, Hanover, New Hampshire, USA; Medical Scientist Training Program, Yale University School of Medicine, New Haven, CT, USA; Department of Anesthesiology, Duke University Medical Center, Durham, North Carolina, USA; Department of Medicine, State University of New York Upstate Medical University, Syracuse, NY, USA; Weill Cornell Medicine, New York, NY USA; Division of Geriatrics and Palliative Medicine, Weill Cornell Medicine, New York, New York, USA; Department of Neurology, University of Texas Southwestern Medical Center, Dallas, TX, USA; O’Donnell Brain Institute, University of Texas Southwestern Medical Center, Dallas, TX USA; Section of Cardiovascular Medicine, Yale School of Medicine, New Haven, CT, USA

**Author notes:** **Corresponding author:** Jennifer M. Kwan, MD, PhD, Yale University School of Medicine New Haven, CT.

## Abstract

**Importance:** Physician-scientist careers depend on sustained research funding, protected time, and institutional resources. However, data on career-retention attitudes across career stages are limited.

**Objective:** To characterize career intentions and associated research-environment factors among MD-PhD students and early-career physician-scientists.

**Design:** Two independent, national cross-sectional surveys were conducted during nonoverlapping periods in 2025; MD-PhD applicant and matriculant trends were also examined.

**Setting:** Medical schools and academic health centers in the United States.

**Participants:** MD-PhD students and early-career physician-scientists (ECPS).

**Exposures:** MD-PhD exposures included perceived institutional support, training-related mental health impact, and research disruption. ECPS exposures included institutional support, adverse funding-related wellbeing, and direct National Institutes of Health funding-process disruption.

**Main Outcome and Measures:** Primary outcomes were consideration of leaving the physician-scientist pathway among MD-PhD students and consideration of leaving academic medicine or research among ECPS. Secondary outcomes included likelihood of staying in academic medicine, research-intensive career preference, and consideration for leaving the US to continue research.

**Results:** Among MD-PhD programs confirmed to have distributed the survey, 838 student responses were received (estimated response rate of 19.3%); 175 ECPS responses were received, with no calculable individual-level response rate. Among MD-PhD students, 423 of 835 (50.7%) reported considering leaving the physician-scientist pathway; 712 of 837 (85.1%) reported a high likelihood of remaining in academic medicine, and 664 of 836 (79.4%) preferred a research-intensive career. Adequate institutional support was associated with lower odds of considering leaving (adjusted odds ratio [aOR], 0.28; 95% CI, 0.14-0.54) and higher odds of staying in academic medicine (aOR, 5.65; 95% CI, 2.91-10.98). Training-related mental health impact (aOR, 2.34; 95% CI, 1.72-3.19) and research disruption (aOR, 1.83; 95% CI, 1.35-2.48) were associated with greater odds of considering leaving. Among ECPS, 57.7% considered leaving, 52.6% reported a high likelihood of staying, and 42.9% considered leaving the United States to continue research. Adverse funding-related well-being was associated with considering leaving academic medicine (aOR, 3.13; 95% CI, 1.52-6.63) and leaving the United States to continue research (aOR, 5.29; 95% CI, 2.35-13.12).

**Conclusions and Relevance:** Consideration of leaving was common despite continued interest in academic and research-intensive careers. Institutional support, adverse well-being, and research disruptions were associated with career intentions.

**Key Points:** *Question:* What research-environment factors are associated with career intentions among MD-PhD trainees and early-career physician-scientists?

*Findings:* In national surveys of 838 MD-PhD students and 175 early-career physician-scientists across the nation, 50.7% and 57.7% respectively reported considering leaving the physician– scientist pathway, although most anticipated remaining in academic medicine. Institutional support was associated with lower odds of considering leaving among MD-PhD students, while adverse well-being was associated with career-exit intentions across career stages.

*Meaning:* Consideration of leaving was common despite continued interest in research-intensive careers. Institutional support, research environment stability, and well-being emerged as potential targets for strengthening the physician-scientist pipeline.

## Introduction

Physician-scientists play a critical role in translating scientific discoveries into advances in patient care. The lengthy physician-scientist training pathway requires institutional investment, mentorship, protected time, and funding across transitions to independence.^1–4^ MD-PhD students and early-career physician-scientists occupy adjacent stages of this pipeline, from entry into physician-scientist training to the transition towards independent research careers.

Instability at either stage may have critical downstream consequences for the future biomedical research workforce. Although most MD-PhD graduates enter positions that may be compatible with research, the amount of time they ultimately devote to research varies substantially,^1,5^ and ECPS report substantial consideration of leaving academic medicine despite continued research participation.^6^ Career intentions may therefore indicate pipeline instability and identify potentially modifiable factors associated with research-career persistence.

These career decisions occur within research environments shaped by institutional support, mentorship, funding stability, and opportunities for career development. Against these longstanding challenges, recent instability in the federal biomedical research environment has introduced additional uncertainty, including disruptions to grant review and award processing and shifts in federal research priorities.^7,8^ Recent studies have identified concerns about research-career stability among biomedical trainees and National Institutes of Health (NIH) career-development award recipients.^9,10^ Prior work during the COVID-19 pandemic similarly demonstrated that large-scale disruptions to the research environment can affect physician-scientist productivity, well-being, and career development.^11^ However, less is known about the research-environment factors associated with career intentions across adjacent stages of physician-scientist development or whether consideration of leaving necessarily reflects reduced interest in research versus uncertainty about the environments in which physician-scientists hope to build their future careers.

We characterized national MD-PhD applicant and matriculant trends and analyzed two independent, nationally-distributed cross-sectional surveys of MD-PhD students and ECPS. Primary outcomes were consideration of leaving the physician-scientist pathway among MD-PhD students and consideration of leaving academic medicine or research among ECPS. We also examined stage-specific outcomes, including likelihood of staying in academic medicine, preference for a research-intensive career, and consideration of leaving the United States (US) to continue research. We hypothesized that lower perceived institutional support, adverse training- or funding-related well-being, and research disruption would be associated with greater consideration of a career exit.

## Methods

### Overall Study Design

We analyzed national Association of American Medical Colleges (AAMC) MD-PhD applicant and matriculant trends and conducted two independent national cross-sectional surveys of MD-PhD students and ECPS, assessing career retention attitudes and factors associated with potential departure.

#### National Applicant Data

Annual MD-PhD applicant, matriculant, and sex-specific matriculant totals from 2012 through 2025 were obtained from the AAMC FACTS report, Table B-8. Matriculation rate was calculated as matriculants divided by applicants. Trends were estimated in Python using locally weighted scatterplot smoothing (LOESS), with a smoothing fraction of 0.4 and 10 robustifying iterations. Years with absolute residuals greater than two standard deviations (SDs) from the fitted trend were classified as outliers. Observed values were plotted together with LOESS curves and ±2-SD bands.

#### MD-PhD Student Survey

Following Institutional Review Board (IRB) approval from the State University of New York Upstate Medical University in August 2025 [IRB# 2346838-2], a 27-question survey was distributed between September 2025 through January 2026 to 104 MD-PhD programs listed by the AAMC. Program directors were asked to forward the survey to current students and received two follow-up emails at 2- to 4-week intervals. Participants provided electronic consent, and anonymous, deidentified responses were collected in REDCap. Email addresses for an optional $25 raffle were stored separately from survey responses. Distributing programs were identified through program-director confirmation and institution-linked raffle responses; their enrolled MD-PhD populations comprised the estimated invitation denominator.

#### ECPS Survey

Following Massachusetts General Hospital IRB approval, a SurveyMonkey questionnaire assessing demographics, career support, clinical and research responsibilities, funding, and career challenges, was distributed from February through August 2025. Early-career physician-scientists were defined as research-focused residents or fellows and investigators who recently completed training. Department chairs at 110 U.S. institutions were asked to share it with their early career investigators. Because department chairs distributed the survey, the individual invitation denominator and response rate could not be determined.

### Statistical Analysis

Descriptive statistics summarized respondent characteristics, key research-environment exposures, and career-intention outcomes. For MD-PhD students, the primary outcome was consideration of leaving the physician-scientist pathway since January 2025. Secondary outcomes were high likelihood of remaining in academic medicine and preference for a research-intensive career, defined as anticipating at least 50% of ideal future effort dedicated to research. Each outcome was modeled as a binary variable. Prespecified exposures included perceived institutional support for a research-intensive career, training-related mental health impact, laboratory or research disruption, program type (Medical Scientist Training Program [MSTP] status), and training stage. Institutional support and training stage were modeled categorically, with no support and preclinical training as the references respectively. Mental health impact and research disruption were modeled as binary variables.

For ECPS, the primary outcome was consideration of leaving academic medicine or research. Secondary outcomes were high likelihood of staying in academic medicine or research, defined as a reported likelihood greater than 75%, and consideration of leaving the US to continue research. Three prespecified binary variables were multidomain institutional support, adverse funding-related well-being, and direct NIH funding-process disruption. Multidomain institutional support was defined as support in at least 2 of 3 domains: career-development award applications, salary equivalent to non-research-track peers, and research-protected effort or relative value unit (RVU) incentives. Adverse funding-related wellbeing impact included reported stress, burnout, depression, or loss of motivation. Direct funding-process disruption included a rescinded NIH award or postponed or canceled study section or council review. For descriptive context, current ECPS responses were compared with published estimates from a prior ECPS survey from 2025.^6^

Multivariable logistic regression was used to investigate the associations with each outcome. Model 1 estimated unadjusted associations, Model 2 included the central research-environment variables, and Model 3 adjusted for MSTP status, race and ethnicity, gender identity, and training stage for the MD-PhD survey and career stage, age, race and ethnicity, and gender identity for ECPS. Model 3 estimates are reported in the main analysis, with full Model 1-3 estimates shown in Supplementary Table 1 and 2. Analyses used complete cases with item-specific denominators. Results are reported as odds ratios or adjusted odds ratios with 95% confidence intervals. Statistical analyses were performed in R version 4.6.0.

## Results

### National Matriculation Data

From 2012 to 2025, the annual number of MD-PhD applicants fluctuated without a consistent increase, while the number of matriculants increased (**Figure 1**). The matriculation rate increased through 2022-2024, but declined in 2025 as applicant growth exceeded matriculant growth. Female matriculants increased and matched or exceeded male matriculants starting in 2019. Relative to the LOESS-fitted trends, 2021 was identified as an outlier year for both applicant and matriculant trends, while the 2025 matriculation rate was an outlier below the fitted trend.

**Figure 1:**
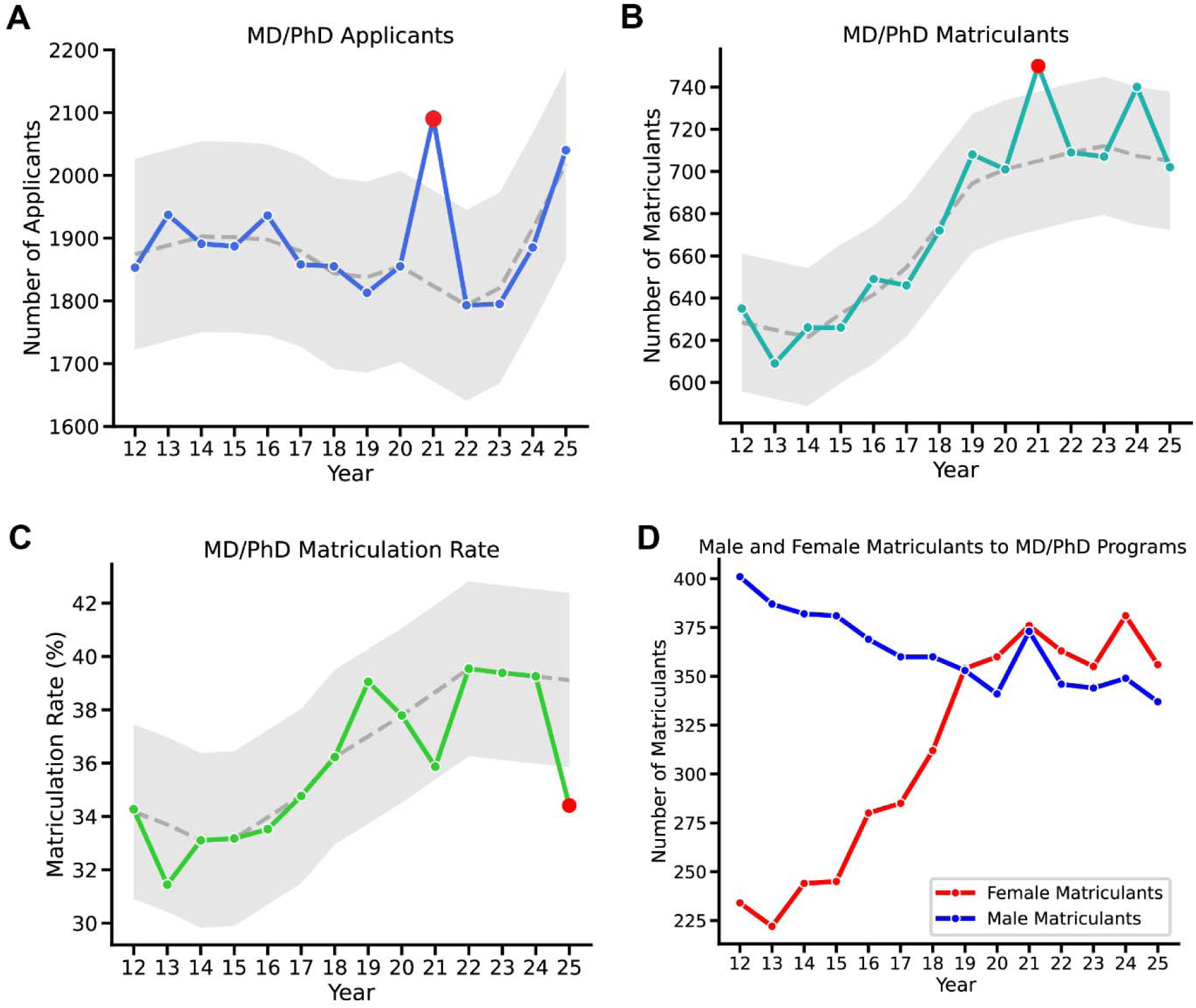
Application and Matriculation Trends for MD-PhD Programs (2012–2025). **A)** Total number of MD-PhD applicants, **B)** total number of MD-PhD matriculants, and **C)** MD-PhD matriculation rate (%) by year. Colored solid lines show observed values; gray dashed lines show LOESS-smoothed trends (smoothing fraction = 0.4, 10 iterations). Shaded regions denote the LOESS confidence band. Red markers indicate outliers, defined as points with residuals exceeding 2 standard deviations (2_σ_) from the predicted LOESS trend. **D)** Number of MD-PhD matriculants by sex (female, red; male, blue) over the same period.

### Survey Respondent Characteristics

The surveys received 916 MD-PhD and 278 ECPS responses, of which 838 and 175, respectively, were submitted (**Table 1**). Among MD-PhD students, 666 of 837 respondents (79.5%) were enrolled in a Medical Scientist Training Program (MSTP), and 450 of 837 respondents (53.8%) were in the graduate PhD phase of training. Among ECPS respondents, 82 of 175 (46.9%) were assistant professors, and 45 (25.7%) were residents or fellows.

**Table 1.** Characteristics of the MD-PhD Trainee and Early-Career Physician-Scientist Surveys.

| Characteristic | MD-PhD Trainee Survey, No. (%) | ECPS Survey, No. (%) |
| --- | --- | --- |
| **Shared demographic characteristics** |  |  |
| **No. of respondents** | 838 | 175 |
| **Age category, No. (%)** |  |  |
| 18-24 | 172 (20.5) | 0 (0.0) |
| 25-34 | 654 (78.0) | 35 (20.0) |
| 35-44 | 11 (1.3) | 106 (60.6) |
| ≥45 | 0 (0.0) | 28 (16.0) |
| **Gender identity, No. (%)** |  |  |
| Female | 431 (51.4) | 78 (44.6) |
| Male | 377 (45.0) | 90 (51.4) |
| Nonbinary, transgender, genderqueer, self-described, or another identity | 29 (3.4) | 4 (2.3) |
| Prefer not to answer / missing | 1 (0.1) | 3 (1.7) |
| **Hispanic, Latino/a, or Spanish origin, No. (%)** |  |  |
| Yes | 50 (6.0) | 10 (5.7) |
| No | 788 (94.0) | 163 (93.1) |
| Prefer not to answer / missing | 0 (0.0) | 2 (1.1) |
| **Race, No. (%)** |  |  |
| White | 424 (50.6) | 113 (64.6) |
| Asian | 229 (27.3) | 36 (20.6) |
| Black or African American | 45 (5.4) | 14 (8.0) |
| American Indian or Alaska Native | 3 (0.4) | 0 (0.0) |
| More than one race | 65 (7.8) | 8 (4.6) |
| Other / self-described | 21 (2.5) | 2 (1.1) |
| Prefer not to answer / missing | 1 (0.1) | 2 (1.1) |
| **Training and professional characteristics** |  |  |
| *MD-PhD trainee survey* |  |  |
| **Program type, No. (%)** |  | *Not collected* |
| MD-PhD, MSTP-funded | 666 (79.5) | - |
| MD-PhD, non-MSTP | 171 (20.4) | - |
| **Training stage, No. (%)** |  | *Not collected* |
| Preclinical medical student | 230 (27.4) | - |
| Graduate/PhD phase | 450 (53.8) | - |
| Clinical medical student | 157 (18.7) | - |
| Research year / other | 0 (0.0) | - |
| *ECPS survey* |  |  |
| **Current position, No. (%)** |  |  |
| Resident | - | 24 (13.7) |
| Fellow | - | 21 (12.0) |
| Instructor | - | 17 (9.7) |
| Assistant professor | - | 82 (46.9) |
| Associate professor | - | 18 (10.3) |
| Professor | - | 8 (4.6) |
| Other | - | 5 (2.9) |
| **Year of terminal training completion, No. (%)** |  |  |
| Prior to 2015 | - | 22 (12.6) |
| 2015-2019 | - | 36 (20.6) |
| 2020-2024 | - | 68 (38.9) |
| Other / not applicable | - | 49 (28.0) |
| **Specialty, No. (%)** |  |  |
| Internal medicine/subspecialties | - | 101 (57.7) |
| Neurology/Psychiatry | - | 18 (10.3) |
| Surgical specialties | - | 8 (4.6) |

| Characteristic | MD-PhD Trainee<br>Survey, No. (%) | ECPS Survey, No. (%) |
| --- | --- | --- |
| Pathology | - | 11 (6.3) |
| Other specialties | - | 37 (21.1) |

Abbreviations: ECPS, early-career physician-scientist.

Among MD-PhD students, 554 of 837 respondents (66.2%) felt adequately supported by their institution in pursuing a research-intensive career, 226 (27.0%) somewhat supported and 48 (5.7%) unsupported (**Table 2**). A training-related mental health impact was reported by 372 of 836 respondents (44.5%), and 370 of 832 respondents (44.5%) reported having experienced at least one lab or research disruption.

**Table 2.** Key Research-Environment Exposures and Career-Intention Outcomes Among Respondents to the MD-PhD Trainee and Early-Career Physician-Scientist Surveys.

| Characteristic | MD-PhD Trainee Survey, No. (%) | ECPS Survey, No. (%) |
| --- | --- | --- |
| **Key research-environment exposures** |  |  |
| *MD-PhD trainee survey* |  |  |
| **Institutional support for research-intensive career** |  | - |
| Yes, adequately supported | 554 (66.2) | - |
| Somewhat supported | 226 (27.0) | - |
| Not supported | 48 (5.7) | - |
| Unsure | 9 (1.1) | - |
| Training-related mental health impact reported | 372 (44.5) | - |
| Any lab/research disruption reported<sup>⊥</sup> | 370 (44.5) | - |
| Grant plans affected by funding changes reported | 66 (7.93) | - |
| *ECPS survey* |  |  |
| Multidomain institutional support reported<sup>⊥</sup> | - | 83 (47.4) |
| Adverse funding-related wellbeing impact reported<sup>⊥</sup> | - | 123 (70.3) |
| Direct NIH funding-process disruption reported<sup>⊥</sup> | - | 40 (22.9) |
| **Career-intention outcomes** |  |  |
| *MD-PhD trainee survey* |  |  |
| Considering leaving the physician-scientist pathway | 423 (50.7) | - |
| High likelihood of staying in academic medicine<sup>⊥</sup> | 712 (85.1) | - |
| Preference for a research-intensive career<sup>⊥</sup> | 664 (79.43) | - |
| *ECPS survey* |  |  |
| Considering leaving academic medicine/research | - | 101 (57.7) |
| High likelihood of staying in academic medicine/research<sup>⊥</sup> | - | 92 (52.6) |
| Considering leaving the United States to continue research | - | 75 (42.9) |

Abbreviations: ECPS, early-career physician-scientist.
⊥ Lab/research disruption (MD-PhD) is marked yes if the respondent indicated any of the following: lab lost funding, stipend reduced/delayed, program/research-year funding lost, or a research opportunity lost/deferred.
⊥ Multidomain institutional support (ECPS) is a composite indicating support across 2-3 domains (career development award support, salary equity between physician-scientists and full-time clinicians, and research incentives/RVUs).
⊥ Adverse funding-related wellbeing impact (ECPS) is coded yes if the respondent reported any of the following: increased stress, burnout, depression symptoms, or loss of motivation in response to funding disruptions.
⊥ Direct NIH funding-process disruption (ECPS) is coded yes if the respondent reported a grant rescinded/paused/delayed and/or a study section or advisory council meeting canceled or postponed.
⊥ High likelihood of staying is defined as a response of > 75% or 100% likelihood of remaining in academic medicine/research within 5 years.
⊥ Preference for a research-intensive career is defined from ideal research/clinical time distribution (e.g., ≥50% research) and/or selection of a research-focused career area.

Among ECPS respondents, 83 (47.4%) reported multidomain institutional support across at least 2 of 3 measured domains (**Table 2**). Adverse funding-related wellbeing impact was reported by 123 individuals (70.3%), and 40 respondents (22.9%) reported having experienced a direct NIH funding-process disruption.

### Career Intentions Among MD-PhD Students

Over half of MD-PhD students (50.7%; 423 of 835 respondents) reported having considered leaving the physician-scientist pathway (**Table 2**). At the same time, 85.1% (712 of 837 respondents) reported a high likelihood of remaining in academic medicine, while 79.4% (664 of 836 respondents) expressed a preference for a research-intensive career. The three most cited reasons that might lead students to consider leaving academic medicine or a research career were: grant/funding challenges (n=711, 84.94%) burnout (n=580, 69.30%), and financial instability (n=577, 68.94%) (**Table 3**).

**Table 3.** Top reasons cited for considering leaving academic medicine or a research-intensive career, by cohort.

|  | No. | % |
| --- | --- | --- |
| ***MD-PhD Trainees*** |  |  |
| Grant/funding challenges | 711 | 84.9 |
| Burnout | 580 | 69.3 |
| Financial instability | 577 | 68.9 |
| Pressure to prioritize clinical productivity | 266 | 31.8 |
| Lack of mentorship | 207 | 24.7 |
| Isolation or lack of peer support | 203 | 24.3 |
| No interest in academia | 97 | 11.6 |
| Other (please specify) | 46 | 5.5 |
| I would not consider leaving | 47 | 5.6 |
| ***Early-Career Physician-Scientists (ECPS)*** |  |  |
| Funding challenges | 129 | 73.7 |
| Undercompensation | 74 | 42.3 |
| Unhappy, stressed, or otherwise less than satisfied with position | 72 | 41.1 |
| Burnout | 66 | 37.7 |
| Drawn to, excited by, or otherwise attracted to a different position | 29 | 16.6 |
| I would not consider leaving | 12 | 6.9 |

In an adjusted logistic regression model, perceived institutional support was strongly associated with career intentions (**Figure 2A; Supplementary Table 1**). Compared with trainees who did not feel supported, trainees who felt adequately supported had lower odds of considering leaving the physician-scientist pathway (adjusted odds ratio [aOR], 0.28; 95% CI, 0.14-0.54), and higher odds of expressing a high likelihood of staying in academic medicine (aOR, 5.65; 95% CI, 2.91-10.98). Adequate support was also associated with a greater likelihood of preferring a research-intensive career (aOR, 2.13; 95% CI, 1.14-4.00). Trainees who felt somewhat supported also had greater odds of reporting a high likelihood of staying in academic medicine (aOR, 3.76; 95% CI, 1.93-7.30) and preferring a research-intensive career (aOR, 2.80; 95% CI, 1.46-5.38).

**Figure 2:**
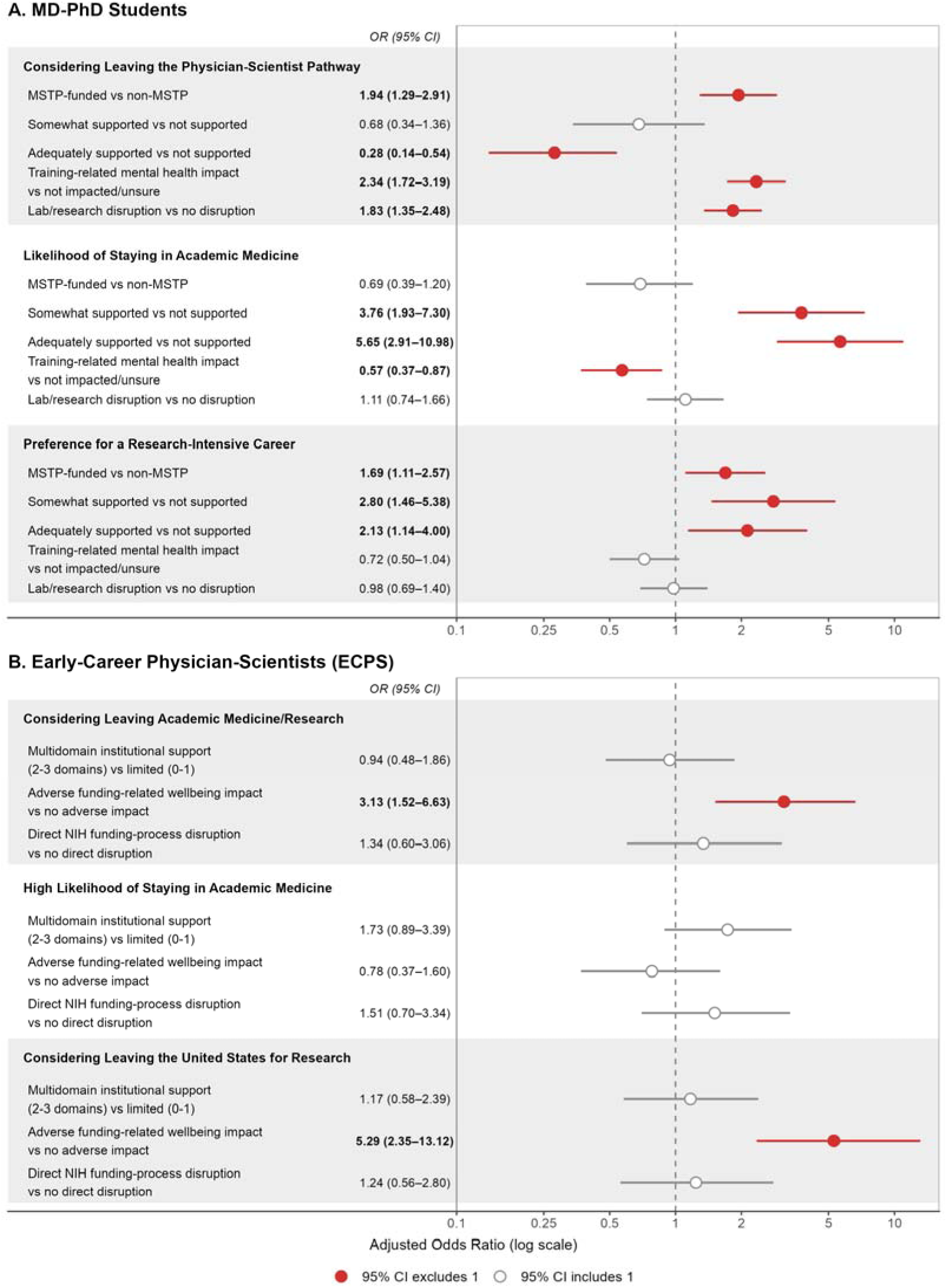
Factors Associated with Career Intentions across the Physician-Scientist Pipeline. Adjusted odds ratios (ORs) and 95% confidence intervals (CIs) are shown for **A)** MD-PhD trainees and **B)** early-career physician-scientists across three career-intention outcomes. Points indicated adjusted ORs, and horizontal lines indicate 95% CIs on a logarithmic scale. The dashed vertical line marks an OR of 1. Red markers demonstrate 95% CIs excluding 1, and empty grey markers indicate 95% CIs including 1.

Reporting a training-related mental health impact was associated with greater odds of considering leaving the physician-scientist pathway (aOR, 2.34; 95% CI, 1.72 - 3.19) and lower odds of reporting a high likelihood of staying in academic medicine (aOR, 0.57; 95% CI, 0.37-0.87). Meanwhile, training-related mental health impact was not significantly associated with preference for a research-intensive career after adjustment (aOR, 0.72; 95% CI, 0.50-1.04).

Lab or research disruption was also associated with considering leaving the physician-scientist pathway (aOR, 1.83; 95% CI, 1.35-2.48) but was not associated with likelihood of staying in academic medicine or preference for a research-intensive career. Compared with preclinical trainees, trainees in the PhD phase (aOR, 1.84; 95% CI, 1.29-2.62) and clinical phase (aOR, 2.16; 95% CI, 1.38-3.40) had greater odds of considering leaving the physician-scientist pathway.

Lastly, enrollment in an MSTP-funded program was associated with greater odds of considering leaving the physician-scientist pathway compared with enrollment in a non-MSTP program (aOR, 1.94; 95% CI, 1.29-2.91). MSTP enrollment was also associated with greater odds of preferring a research-intensive career (aOR, 1.69; 95% CI, 1.11-2.57) but was not associated with likelihood of staying in academic medicine (aOR, 0.69; 95% CI, 0.39-1.20).

### Career Intentions Among Early-Career Physician-Scientists

Of ECPS respondents, over half (57.7%) reported considering leaving academic medicine or research, 52.6% reported a high likelihood of staying in academic medicine, and 42.9% reported considering leaving the US to continue research (**Table 2**). Just over half of ECPS (50.8%) reported having received a career development award. However, the proportion considering leaving academic medicine or research did not differ significantly between individuals with and without a career development award (58.9% vs. 50.6%; p = 0.37).

Consideration of leaving the US to continue research was also similar between these groups (45.6% vs. 38.8%; p = 0.43). The top three reasons that might lead an ECPS to leave their position were funding challenges (n=129, 73.71%), undercompensation (n=74, 42.29%), and feeling “unhappy, stressed, or otherwise less than satisfied with my current position” (n=72, 41.14%) **(Table 3)**. A descriptive comparison with the prior ECPS cohort is shown in **Supplementary Figure 1**.

In an adjusted logistic regression model, adverse funding-related wellbeing impact was associated with greater odds of considering leaving academic medicine or research (aOR, 3.13; 95% CI, 1.52-6.63) (**Figure 2B; Supplementary Table 2**). Adverse funding-related wellbeing impact was also associated with greater odds of considering leaving the US to continue research (aOR, 5.29; 95% CI, 2.35-13.12). It was not associated with a high likelihood of staying in academic medicine or research (aOR, 0.78; 95% CI, 0.37-1.60).

On the other hand, multi-domain institutional support was not independently associated with considering leaving academic medicine or research (aOR, 0.94; 95% CI, 0.48-1.86), high likelihood of staying in academic medicine or research (aOR, 1.73; 95% CI, 0.89-3.30), or considering leaving the US to continue research (aOR, 1.17; 95% CI, 0.58-2.39). Similarly, direct NIH funding-process disruption was not independently associated with any of the ECPS career-intention outcomes.

## Discussion

In these two national surveys, consideration of leaving the physician-scientist pathway was common among both MD-PhD students and ECPS, although most respondents anticipated remaining in academic medicine or pursuing research-intensive careers. Among MD-PhD students, perceived institutional support was associated with lower odds of considering leaving, while training-related mental health and laboratory or research disruption were associated with higher odds. Among ECPS, adverse funding-related wellbeing was associated with considering leaving academic medicine or research and leaving the US to continue research.

The apparent disconnect between career-exit consideration and continued interest in academic medicine and research is an important finding. Among MD-PhD students, training-related mental health impact was associated with considering leaving and lower likelihood of remaining in academic medicine, but not with preference for a research-intensive career.

Similarly, 42.9% of ECPS considered leaving the US specifically to continue research. These findings suggest that considering departure does not necessarily indicate loss of scientific motivation. For some respondents, it may instead reflect uncertainty about whether their current training, funding, or research environments can support a sustainable research career.

National AAMC trends provide complementary context: MD-PhD applicant numbers remained relatively stable from 2012 through 2025 while matriculant numbers increased. However, the high prevalence of career-exit consideration among current trainees suggests that maintaining the physician-scientist pipeline depends not only on attracting and enrolling students, but also on sustaining their confidence in the pathway after matriculation. MD-PhD programs have traditionally been highly successful in retaining trainees through dual-degree completion.^6,12^ However, the transition from clinical training to an independent research career remains one of the most vulnerable points in the physician-scientist pipeline with a high level of attrition, when investigators must secure protected research time and sustained funding. National physician-scientist leaders have identified the transition from trainee to independent investigator as among the most pressing unmet needs in the career pathway.^13^ However, the current research environment appears to threaten even the MD-PhD training stage and worsen the already existent threat^6^ to early career investigators. Together, these findings highlight retention and training-environment stability as important components to recruitment.

Among MD-PhD students, perceived institutional support had the most consistent association with career intentions. Adequate support was associated with lower odds of considering leaving and greater odds of anticipating academic and research-intensive careers. These data are consistent with prior work linking mentorship and supportive institutional environments with career persistence and well-being.^14,15^ Given that perceived support and career intentions were measured concurrently, these associations cannot establish whether institutional support itself reduces career-exit consideration; however, they identify perceived institutional support as a potentially important correlate of career intentions. The consistency across outcomes further suggests that institutional commitment to research-intensive careers may be an important marker of confidence in the physician-scientist pathway.

Training-related mental health impact and research disruption were also associated with considering leaving, while PhD- and clinical-phase trainees had higher odds than preclinical trainees, identifying possible periods of vulnerability. Unexpectedly, MSTP enrollment was associated with both greater odds of considering leaving and stronger preference for a research-intensive career. This differs from prior findings that MSTP graduates are more likely to obtain full-time faculty appointments and that attrition is higher at non-MSTP programs.^5,16^ This finding should be explored further, particularly in the context of recent federal funding changes that may have created more uncertainty for MSTP-supported programs than for programs relying on other funding sources.

Among ECPS, adverse funding-related well-being was associated with both career-exit consideration and consideration of leaving the US to continue research. This finding extends previous evidence that burnout and lack of professional fulfillment are associated with intention to leave academic medicine among academic physicians.^17,18^ Similarly, recent data investigating NIH K awardees identified widespread concern about the stability of research careers following federal policy changes.^9^ Direct NIH funding-process disruption, reported by 22.9% of respondents, was not independently associated with these outcomes. Interestingly, just over half of ECPS respondents had received a career development award, however, career departure and relocation considerations were similarly prevalent among physician-scientists with and without an award. Multidomain institutional support was also not independently associated with ECPS career intentions. These results do not indicate that institutional support is unimportant. Rather, the support pillars measured in this survey, including career-development award assistance, salary equity, and protected research efforts, may not capture the scale or duration of resources needed to establish an independent research program. Therefore, the relationship between institutional support and career intentions may differ between trainees and investigators transitioning to independence.

These findings have stage-specific implications for institutional and federal stakeholders. MD-PhD programs can support retention through clear communication about funding and training continuity, offer support for students affected by laboratory disruptions, stage-specific advising, and access to mental health resources. Academic medical centers can protect research effort, provide bridge and grant-development support, and align salary and promotion structures to support early-career investigators. Since early-career independence remains closely tied to external federal research funding, predictable grant-review and funding processes are also central to maintaining a stable physician-scientist workforce.

## Limitations

This study had several limitations. First, the cross-sectional design of the surveys prevents any causal inferences between whether reported career intentions will translate into actual career changes or if perceptions have changed over time. Moreover, the MD-PhD and ECPS surveys were distributed over adjacent but nonoverlapping periods and used different instruments and outcome definitions, preventing direct statistical comparison between cohorts. Exposures and outcomes were self-reported and measured concurrently, creating potential common-method bias and reverse causation. Several constructs were measured using individual survey items rather than validated multidimensional instruments. Furthermore, the relatively small ECPS sample and inability to determine an individual-level response rate introduce potential selection and nonresponse bias.

## Conclusion

Across adjacent stages of the physician-scientist pipeline, consideration of leaving was common despite continued interest in academic and research-intensive careers. Institutional support, well-being, research disruption, and training stage showed differing associations with career intentions. These data suggest that instability in the physician-scientist pathway may reflect uncertainty about whether current institutional, training, and funding environments can support this scientific motivation, rather than loss of scientific commitment alone. Longitudinal studies are needed to determine whether these intentions predict subsequent research participation, geographic relocation, or departure from the physician-scientist workforce.

## Data Availability

All data produced in this study may be made available upon reasonable request to the corresponding author, subject to institutional review board requirements.

## Acknowledgements

We thank the American Junior Investigator Association for its support of this study. We also thank the physician-scientist training program directors and coordinators who assisted with survey-distribution and all participants who shared their experiences.

