## Supplemental Tables and Figure for "Career Intentions and Research-Environment Factors Across the Physician-Scientist Pipeline"

| **Table 1. Training-Related Factors Associated with MD-PhD Trainee Career Intentions** | | | | | | | | | |
| --- | --- | --- | --- | --- | --- | --- | --- | --- | --- |
| **Characteristic** | **Considering Leaving Physician-Scientist Pathway** | | | **Likelihood of Staying in Academic Medicine** | | | **Preference for Research-Intensive Career** | | |
|  | **Model 1: Unadjusted OR (95% CI)** | **Model 2: Adjusted OR without Demographics** | **Model 3: Adjusted OR with Demographics** | **Model 1: Unadjusted OR (95% CI)** | **Model 2: Adjusted OR without Demographics** | **Model 3: Adjusted OR with Demographics** | **Model 1: Unadjusted OR (95% CI)** | **Model 2: Adjusted OR without Demographics** | **Model 3: Adjusted OR with Demographics** |
| No. of participants | 831-835 | 831 | 831 | 832-837 | 832 | 832 | 832-836 | 832 | 832 |
| **Program type (ref: Non-MSTP)** | | | | | | | | | |
| Non-MSTP MD-PhD | *Ref* | *Ref* | *Ref* | *Ref* | *Ref* | *Ref* | *Ref* | *Ref* | *Ref* |
| MSTP-funded MD-PhD | 1.27 (0.91, 1.78) | **1.93 (1.30, 2.86)** | **1.94 (1.29, 2.91)** | 1.03 (0.64, 1.64) | 0.74 (0.43, 1.27) | 0.69 (0.39, 1.20) | **1.79 (1.22, 2.63)** | **1.65 (1.09, 2.48)** | **1.69 (1.11, 2.57)** |
| **Institutional support (ref: not supported)** | | | | | | | | | |
| Not supported | *Ref* | *Ref* | *Ref* | *Ref* | *Ref* | *Ref* | *Ref* | *Ref* | *Ref* |
| Somewhat supported | 0.73 (0.38, 1.40) | 0.72 (0.37, 1.41) | 0.68 (0.34, 1.36) | **3.24 (1.72, 6.10)** | **3.24 (1.69, 6.19)** | **3.76 (1.93, 7.30)** | **2.88 (1.54, 5.38)** | **2.54 (1.34, 4.82)** | **2.80 (1.46, 5.38)** |
| Adequately supported | **0.25 (0.14, 0.47)** | **0.28 (0.15, 0.54)** | **0.28 (0.14, 0.54)** | **5.27 (2.92, 9.52)** | **4.78 (2.51, 9.11)** | **5.65 (2.91, 10.98)** | **2.85 (1.61, 5.05)** | **2.07 (1.12, 3.83)** | **2.13 (1.14, 4.00)** |
| **Training-related mental health impact (ref: not impacted)** | | | | | | | | | |
| Not impacted/Unsure | *Ref* | *Ref* | *Ref* | *Ref* | *Ref* | *Ref* | *Ref* | *Ref* | *Ref* |
| Impacted | **3.18 (2.39, 4.23)** | **2.66 (1.97, 3.59)** | **2.34 (1.72, 3.19)** | **0.44 (0.30, 0.65)** | **0.52 (0.35, 0.79)** | **0.57 (0.37, 0.87)** | **0.60 (0.43, 0.84)** | **0.63 (0.44, 0.89)** | 0.72 (0.50, 1.04) |
| **Lab/research disruption (ref: no disruption)** | | | | | | | | | |
| No disruption | *Ref* | *Ref* | *Ref* | *Ref* | *Ref* | *Ref* | *Ref* | *Ref* | *Ref* |
| Disruption experienced | **2.04 (1.54, 2.69)** | **1.84 (1.37, 2.48)** | **1.83 (1.35, 2.48)** | 0.88 (0.60, 1.29) | 1.01 (0.68, 1.50) | 1.11 (0.74, 1.66) | 0.90 (0.64, 1.26) | 0.96 (0.68, 1.36) | 0.98 (0.69, 1.40) |
| ***Demographic Variables (included in Model 3 only)*** | | | | | | | | | |
| **Race/ethnicity (ref: Non-URM)** | | | | | | | | | |
| Non-URM | *Ref* | - | *Ref* | *Ref* | - | *Ref* | *Ref* | - | *Ref* |
| URM | 1.02 (0.74, 1.42) | - | 1.11 (0.77, 1.59) | 0.79 (0.51, 1.22) | - | 0.71 (0.45, 1.11) | 1.04 (0.69, 1.56) | - | 0.93 (0.61, 1.42) |
| **Gender identity (ref: Male)** | | | | | | | | | |
| Male | *Ref* | - | *Ref* | *Ref* | - | *Ref* | *Ref* | - | *Ref* |
| Female | 1.23 (0.94, 1.61) | - | 1.13 (0.84, 1.54) | 0.78 (0.53, 1.14) | - | 0.73 (0.49, 1.11) | **0.71 (0.51, 1.00)** | - | **0.68 (0.47, 0.98)** |
| Other/Non-binary | 0.91 (0.43, 1.90) | - | 0.86 (0.37, 1.96) | 0.84 (0.31, 2.24) | - | 0.86 (0.29, 2.55) | 1.25 (0.47, 3.33) | - | 1.09 (0.38, 3.13) |
| **Training stage (ref: Pre-clinical)** | | | | | | | | | |
| Pre-clinical (M1, M2) | *Ref* | - | *Ref* | *Ref* | - | *Ref* | *Ref* | - | *Ref* |
| PhD | **1.59 (1.21, 2.10)** | - | **1.84 (1.29, 2.62)** | **0.52 (0.35, 0.77)** | - | **0.59 (0.35, 0.99)** | 0.78 (0.55, 1.09) | - | **0.58 (0.37, 0.93)** |
| Clinical (M3, M4) | **1.61 (1.13, 2.30)** | - | **2.16 (1.38, 3.40)** | 1.44 (0.85, 2.46) | - | 1.24 (0.62, 2.49) | 0.68 (0.45, 1.01) | - | **0.45 (0.26, 0.78)** |
| *Abbreviations: CI = Confidence Interval, OR = Odds Ratio; URM= Underrepresented Minority. Model 1 = unadjusted/univariate logistic regression. Model 2 = adjusted excluding demographic covariates. Model 3 = adjusted including demographic covariates: race/ethnicity, gender identity, training stage, and program type. Research-intensive career intentions were defined as reporting that ≥50% of ideal future career time would be spent in research, compared with <50%. Bold values indicate statistical significance (p < 0.05). "-" indicates variable not included in that model. N varies across univariate models due to missing responses in individual predictors.* | | | | | | | | | |

| **Table 2. Funding- and Support-Related Factors Associated with Early-Career Physician-Scientist (ECPS) Career Intentions** | | | | | | | | | |
| --- | --- | --- | --- | --- | --- | --- | --- | --- | --- |
| **Characteristic** | **Considering Leaving Academic Medicine/Research** | | | **High Likelihood of Staying in Academic Medicine/Research** | | | **Considering Leaving the United States to Continue Research** | | |
|  | **Model 1: Unadjusted OR (95% CI)** | **Model 2: Adjusted OR without Demographics** | **Model 3: Adjusted OR with Demographics** | **Model 1: Unadjusted OR (95% CI)** | **Model 2: Adjusted OR without Demographics** | **Model 3: Adjusted OR with Demographics** | **Model 1: Unadjusted OR (95% CI)** | **Model 2: Adjusted OR without Demographics** | **Model 3: Adjusted OR with Demographics** |
| No. of participants | 169–175 | 171 | 162 | 169–175 | 171 | 162 | 167–175 | 169 | 160 |
| **Multidomain institutional support (ref: limited support, 0–1 domains)** | | | | | | | | | |
| Limited support (0–1 domains) | *Ref* | *Ref* | *Ref* | *Ref* | *Ref* | *Ref* | *Ref* | *Ref* | *Ref* |
| Multidomain support (2–3 domains) | 1.03 (0.56, 1.88) | 0.96 (0.51, 1.82) | 0.94 (0.48, 1.86) | **1.94 (1.06, 3.56)** | 1.84 (0.99, 3.42) | 1.73 (0.89, 3.39) | 1.02 (0.56, 1.87) | 1.00 (0.51, 1.96) | 1.17 (0.58, 2.39) |
| **Adverse funding-related wellbeing impact (ref: no adverse impact)** | | | | | | | | | |
| No adverse wellbeing impact | *Ref* | *Ref* | *Ref* | *Ref* | *Ref* | *Ref* | *Ref* | *Ref* | *Ref* |
| Adverse wellbeing impact | **3.05 (1.54, 6.04)** | **2.85 (1.41, 5.73)** | **3.13 (1.52, 6.63)** | 0.96 (0.49, 1.86) | 0.90 (0.46, 1.77) | 0.78 (0.37, 1.60) | **6.36 (2.75, 14.70)** | **6.28 (2.67, 14.73)** | **5.29 (2.35, 13.12)** |
| **Direct NIH funding-process disruption (ref: no disruption)** | | | | | | | | | |
| No direct disruption | *Ref* | *Ref* | *Ref* | *Ref* | *Ref* | *Ref* | *Ref* | *Ref* | *Ref* |
| Direct disruption experienced | 1.71 (0.81, 3.60) | 1.39 (0.65, 2.97) | 1.34 (0.60, 3.06) | 1.48 (0.72, 3.03) | 1.33 (0.63, 2.81) | 1.51 (0.70, 3.34) | 1.37 (0.68, 2.79) | 0.99 (0.45, 2.18) | 1.24 (0.56, 2.80) |
| ***Demographic Variables (included in Model 3 only)*** | | | | | | | | | |
| **Career stage (ref: Resident/Fellow)** | | | | | | | | | |
| Resident/Fellow | *Ref* | *Ref* | *Ref* | *Ref* | *Ref* | *Ref* | *Ref* | *Ref* | *Ref* |
| Instructor/Faculty | 1.23 (0.62, 2.44) | — | 1.00 (0.40, 2.47) | 0.92 (0.47, 1.83) | — | 0.66 (0.26, 1.61) | 0.58 (0.29, 1.16) | — | 0.50 (0.18, 1.30) |
| Other † (n=5; imprecise) | 3.50 (0.36, 33.82) | — | 3.58 (0.48, 43.66) | NE † | — | **0.07 (0.00, 0.83)** | 0.21 (0.02, 2.02) | — | 0.41 (0.03, 3.30) |
| **Age (ref: 25–34 years)** | | | | | | | | | |
| Age 25–34 years | *Ref* | *Ref* | *Ref* | *Ref* | *Ref* | *Ref* | *Ref* | *Ref* | *Ref* |
| Age ≥35 years | 1.36 (0.64, 2.86) | — | 1.12 (0.42, 2.99) | 1.03 (0.49, 2.17) | — | 1.15 (0.44, 3.04) | 0.75 (0.35, 1.60) | — | 0.95 (0.33, 2.75) |
| **Race/ethnicity (ref: Non-URM)** | | | | | | | | | |
| Non-URM | *Ref* | *Ref* | *Ref* | *Ref* | *Ref* | *Ref* | *Ref* | *Ref* | *Ref* |
| URM | 1.56 (0.59, 4.08) | — | 1.34 (0.51, 3.81) | 0.49 (0.19, 1.26) | — | 0.55 (0.20, 1.42) | 0.80 (0.31, 2.08) | — | 0.96 (0.34, 2.64) |
| Multiracial ‡ (n=8; imprecise) | 1.30 (0.30, 5.63) | — | 1.19 (0.25, 6.20) | 0.48 (0.11, 2.08) | — | 0.48 (0.10, 2.16) | 0.72 (0.17, 3.13) | — | 0.43 (0.07, 2.09) |
| **Gender identity (ref: Man)** | | | | | | | | | |
| Man | *Ref* | *Ref* | *Ref* | *Ref* | *Ref* | *Ref* | *Ref* | *Ref* | *Ref* |
| Woman | 1.05 (0.57, 1.94) | — | 1.10 (0.57, 2.13) | 0.92 (0.50, 1.69) | — | 0.97 (0.51, 1.85) | 0.92 (0.50, 1.70) | — | 0.91 (0.46, 1.78) |
| Gender-diverse | 0.73 (0.10, 5.42) | — | 0.36 (0.03, 3.00) | 2.62 (0.26, 26.20) | — | 1.83 (0.22, 23.44) | 3.67 (0.37, 36.71) | — | 1.33 (0.15, 17.01) |
| *Abbreviations: CI = confidence interval, ECPS = early-career physician-scientist, OR = odds ratio, URM = underrepresented minority. Model 1 = unadjusted/univariate logistic regression. Model 2 = adjusted for the three core constructs (institutional support, wellbeing impact, funding-process disruption) without demographic covariates. Model 3 = Model 2 additionally adjusted for career stage, age, race/ethnicity (URM status), and gender identity.* | | | | | | | | | |

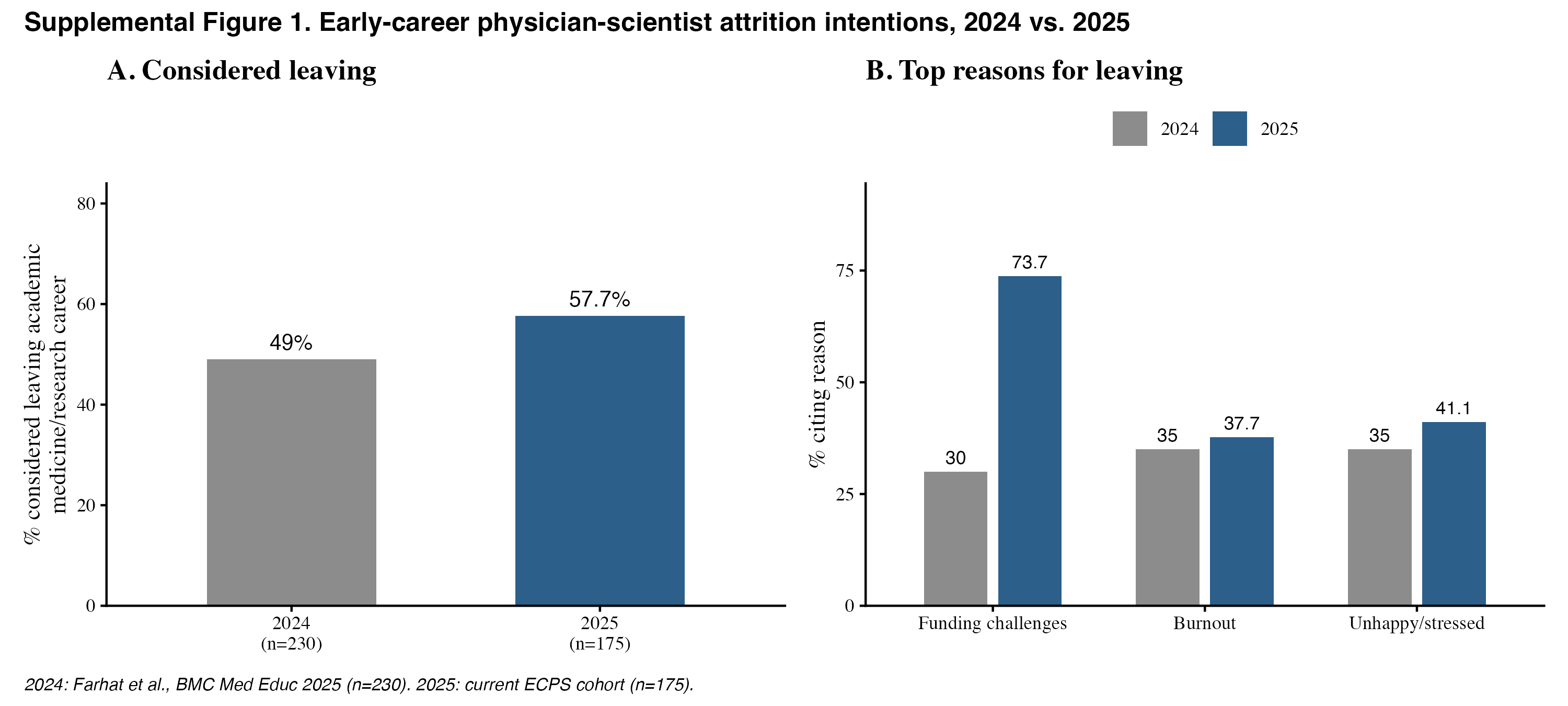
